# Opportunistic Screening of Chronic Liver Disease with Deep Learning Enhanced Echocardiography

**DOI:** 10.1101/2024.06.13.24308898

**Authors:** Yuki Sahashi, Milos Vukadinovic, Fatemeh Amrollahi, Hirsh Trivedi, Justin Rhee, Jonathan Chen, Susan Cheng, David Ouyang, Alan C. Kwan

**Affiliations:** Department of Cardiology, Smidt Heart Institute, Cedars-Sinai Medical Center, Los Angeles, CA; Department of Bioengineering, University of California Los Angeles, Los Angeles, CA; Bioinformatics Research, Department of Medicine, Stanford University, Palo Alto, CA; Karsh Division of Gastroenterology and Hepatology, Department of Medicine, Cedars-Sinai Medical Center, Los Angeles, CA; School of Medicine, Brown University, Providence, RI; Division of Artificial Intelligence in Medicine, Cedars-Sinai Medical Center, Los Angeles, CA

**Author notes:** Correspondence, 127 S. San Vicente Blvd, AHSP A3600 Los Angeles, CA 90048.

**Keywords:** Opportunistic screening, Echocardiography, Deep Learning, Convolutional neural network, Cirrhosis, Steatotic Liver Disease, Non-Alcoholic Fatty Liver Disease

## Abstract

**Importance:** Chronic liver disease affects more than 1.5 billion adults worldwide, however the majority of cases are asymptomatic and undiagnosed. Echocardiography is broadly performed and visualizes the liver; but this information is not leveraged.

**Objective:** To develop and evaluate a deep learning algorithm on echocardiography videos to enable opportunistic screening for chronic liver disease.

**Design:** Retrospective observational cohorts

**Setting:** Two large urban academic medical centers

**Participants:** Adult patients who received echocardiography and abdominal imaging (either abdominal ultrasound or abdominal magnetic resonance imaging) with ≤30 days between tests, between July 4, 2012, to June 4, 2022.

**Exposure:** Deep learning model predictions from a deep-learning computer vision pipeline that identifies subcostal view echocardiogram videos and detects the presence of cirrhosis or steatotic liver disease (SLD).

**Main Outcome and Measures:** Clinical diagnosis by paired abdominal ultrasound or magnetic resonance imaging (MRI).

**Results:** A total of 1,596,640 echocardiogram videos (66,922 studies from 24,276 patients) from Cedars-Sinai Medical Center (CSMC) were used to develop EchoNet-Liver, an automated pipeline that identifies high quality subcostal images from echocardiogram studies and detects the presence of cirrhosis or SLD. In the held-out CSMC test cohort, EchoNet-Liver was able to detect the presence of cirrhosis with an AUC of 0.837 (0.789 - 0.880) and SLD with an AUC of 0.799 (0.758 - 0.837). In a separate test cohort with paired abdominal MRIs, cirrhosis was detected with an AUC of 0.704 (0.689-0.718) and SLD was detected with an AUC of 0.726 (0.659-0.790). In an external test cohort of 106 patients (n = 5,280 videos), the model detected cirrhosis with an AUC of 0.830 (0.738 - 0.909) and SLD with an AUC of 0.768 (0.652 – 0.875).

**Conclusions and Relevance:** Deep learning assessment of clinical echocardiography enables opportunistic screening of SLD and cirrhosis. Application of this algorithm may identify patients who may benefit from further diagnostic testing and treatment for chronic liver disease.

**KEY POINTS:** *Question:* Can a deep learning algorithm applied to echocardiography videos effectively identify chronic liver diseases including cirrhosis and steatotic liver disease (SLD)?

*Findings:* This retrospective observational cohort study utilized 1,596,640 echocardiography videos from 66,922 studies of 24,276 patients. The deep learning model with a computer vision pipeline (EchoNet-Liver) demonstrated strong performance to detect cirrhosis and SLD. External validation at a geographically distinct site demonstrated similar discriminative ability.

*Meaning:* The application of EchoNet-Liver to echocardiography could aid opportunistic screening of chronic liver diseases, providing a unique cost-effective angle to improve patient management.

## Introduction

Chronic liver disease (CLD) affects an estimated 1.5 billion people worldwide and 100 million in the United States and can result in malignancy, end-stage liver disease, or mortality^1^. The prevalence of chronic liver disease is sharply increasing^2,3^, particularly related to steatotic liver disease (SLD) as a result of increased burden of obesity and metabolic disease, however the vast majority of patients are undiagnosed^4,5^. This issue affects many individuals with known cardiovascular disease as well as individuals with even severe end-stage liver disease, such as cirrhosis, which is frequently missed in the earlier stages of fibrosis^4^.

Multiple approaches are available for screening and diagnosis of chronic liver disease, including serological risk scores, qualitative and quantitative ultrasound and magnetic resonance imaging (MRI), and invasive biopsy^6,7^. However, accuracy, availability, and cost all limit the sufficiency of these pathways to address underdiagnosis. Echocardiography, or ultrasound of the heart and associated structures, is a first-line diagnostic ultrasound test, and is frequently performed across the spectrum of patients with metabolic and cardiovascular diseases^8^. Included within a standard echocardiographic examination are subcostal views which visualize the inferior vena cava and provide clear visualization of hepatic tissue quality as well as liver contour. The full clinical utility of these images for identification of hepatic disease is underutilized as cardiologists are not trained in the assessment of liver pathologies by ultrasound.

Artificial intelligence (AI) can identify diseases and characteristics that may not be readily observable by the human eye^9–13^, predict disease progression^14^, mortality^15^, and improve measurement accuracy of cardiac parameters^16–19^. Our study aims to develop and validate an AI computer vision approach to leverage echocardiographic images and videos to detect CLD. We hypothesize that a deep-learning pipeline can identify high-quality subcostal view videos and detect SLD and cirrhosis in a high-throughput fashion. We trained and evaluated the model performance in internal and external validations across multiple cohorts. AI-based analysis of hepatic tissue visible within standard subcostal echocardiography videos may provide opportunistic screening of chronic liver disease from standard echocardiography without additional costs.

## Methods

### Cohort Selection

We included adult patients over 18 years who received an echocardiogram at Cedars-Sinai Medical Center (CSMC) within 30 days of abdominal ultrasound and no history of liver transplant between July 2012 and June 2022. Clinical diagnoses from the abdominal ultrasound report including normal liver, steatotic liver, and cirrhotic liver, were paired with the echocardiogram images as labels for training and validation. In addition, we created an independent test cohort of patients with echocardiography and a matched abdominal MRI for cross-modal validation. Studies from patients included within any test cohort (ultrasound or MRI) were excluded from training cohort (**Supplementary Tables 1 - 4**). A cohort of patients from Stanford Healthcare (SHC) who had echocardiography and abdominal ultrasound within 30 days were included as external validation. An overall flowchart for model development and evaluation is provided in **Figure 1**. Approval for this study was obtained from the Cedars-Sinai Medical Center and Stanford Healthcare Institutional Review Boards, and the requirement for informed consent was waived for retrospective data analysis without patient contact.

**Figure 1:**
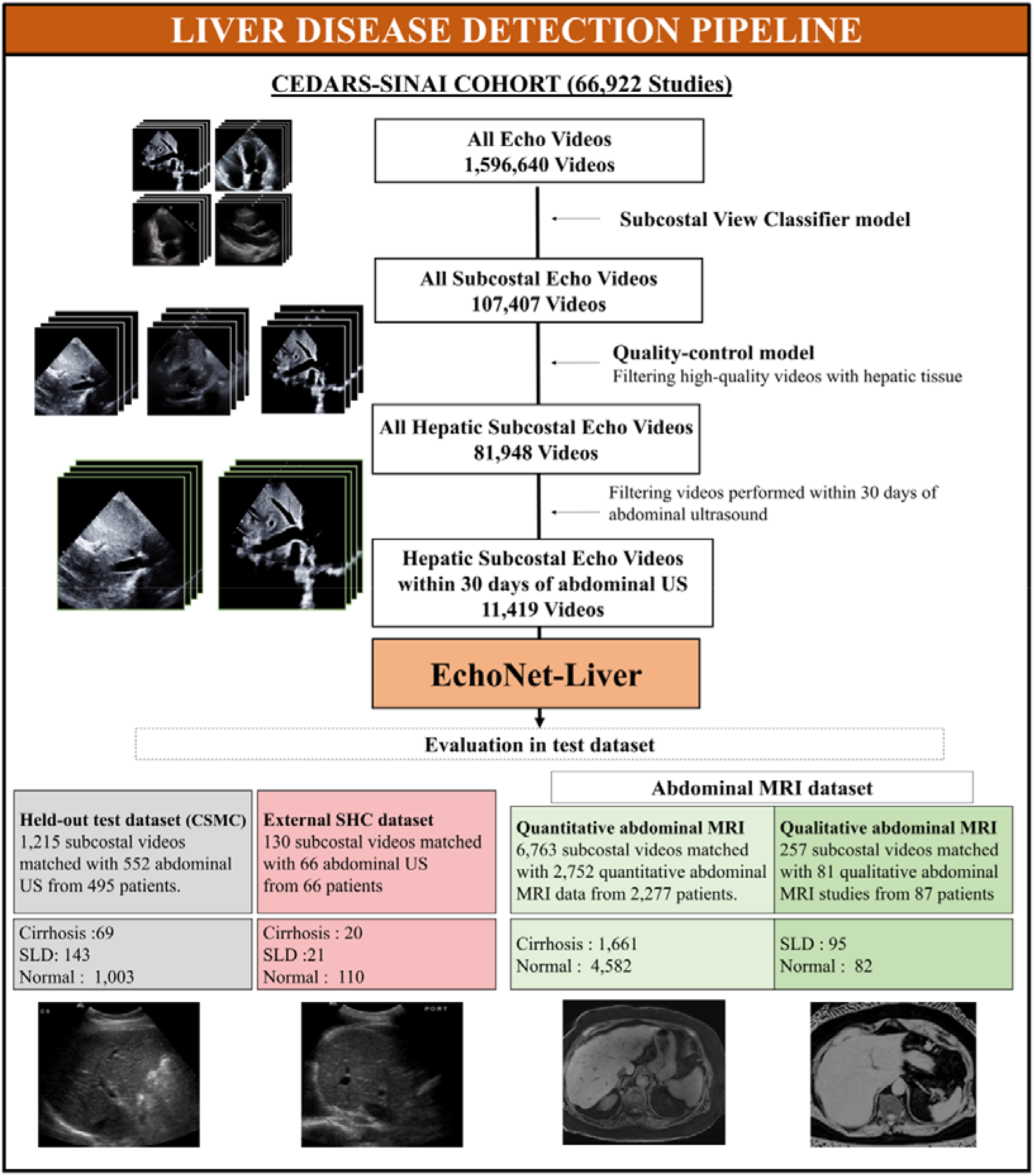
Overview of the study pipeline. More than 2M echocardiogram videos were used to train EchoNet-Liver, an automated pipeline for deep learning view classification, image quality assessment, and detection of chronic liver disease. Evaluation of EchoNet-Liver was performed 3 held-out test cohorts, including patients with paired echocardiograms and abdominal ultrasounds (CSMC), abdominal MRI (CSMC), and an ultrasound external test dataset (SHC). SLD: steatotic liver disease, MRI: magnetic resonance imaging.

### Disease Definitions

Echocardiography studies were matched to the closest abdominal ultrasound study within 30 days. CLD labels were derived from the abdominal ultrasound reports and categorized studies into normal, cirrhosis, or SLD based on text describing the liver parenchyma. If an echocardiography study had greater than one abdominal ultrasound within 30 days, labels were taken from the radiologist confirmed clinical report closest in time. Controls were identified when the report specified the liver parenchyma was normal In the abdominal MRI group, diagnosis labels for cirrhosis was taken from the clinical MRI report; for SLD, we focused on the subset of cases where magnetic resonance imaging-derived proton density fat fraction (MRI-PDFF) or magnetic resonance spectroscopy (MRS) fat signal was measured, with larger than 5.0% being diagnostic of SLD^25^.

### View Selection and Image Processing

A standard echocardiogram study often contains 50-100 videos, of which typically only 1-2 videos per study capture the liver in sufficient quality for assessment of the liver echotexture and contour. Echocardiography videos were initially obtained as Digital Imaging and Communications in Medicine (DICOM) files and underwent de-identification and processing into AVI videos. We developed a pipeline of two deep learning models for view classification (identification of the subcostal view videos) and quality control (excluding videos with severe motion artifact and low image quality). Two video-based convolutional neural networks (R2+1D)^23^ were used with standardized 112 × 112 pixel videos for input. A dataset of 11,778 manually curated videos of 4,991 patients was used model training, classified by both view type and image quality (representative ground truth images in **Supplemental Figure 1**). To evaluate the performance of view selection and quality checking, we randomly selected 100 CSMC echocardiogram studies (n = 2,315 videos) and two cardiologists manually identified high quality subcostal and low-quality subcostal view videos for comparison with model output.

### Chronic Liver Disease Detection

For the training and evaluation of the liver disease detection models, we utilized an image-based model (DenseNet^26^), to focus on hepatic tissue texture for the prediction of cirrhosis and SLD. Given the focus on texture, the input data were still frame images from high quality subcostal echocardiogram videos at native resolution (480 × 640 pixels). We trained the model to minimize binary cross-entropy loss using an AdamW optimizer with an initial learning rate of 1×10^-5^ at a batch size of 40 for 100 epochs. A variety of hyperparameters and architectures were compared prior to development of the final EchoNet-Liver model (**Supplemental Table 5**). All model training and evaluation were conducted using Python 3.8, PyTorch 2.2, and torchvision 0.17.

### Statistical analysis

Model performance was determined by measuring the area under the receiver operating curve (AUC) on held out test cohorts. Area under the precision-recall curves (AUPRC), sensitivity, specificity, positive predictive value, and negative predictive value were similarly reported at the Youden index. All 95% confidence intervals were calculated with 10,000 bootstrapping samples. Data analysis was performed using both Python (version 3.8.0) and R (version 4.2.2) programming languages. This study was carried out following the TRIPOD-AI guideline (**Supplemental Material 1**)^27^. Saliency maps were generated to identify the areas of interest for the classifier across all test datasets. Each saliency map was produced using Grad-CAM^28^, which captures the gradient information directed into the final convolutional layer of the trained deep learning model. We input the final layer of the fourth DenseBlock for this approach.

### Code and Data Availability

Code is available at https://github.com/echonet/liver/ and training data is available with submission of a research protocol and approval by CSMC and SHC IRBs.

## RESULTS

### Patient characteristics

Using a total of 1,596,640 videos from 66,922 CSMC echocardiography studies of 24,276 patients were identified and split 8:1:1 by patient into training, validation, and held-out test cohorts. The patient cohorts exhibited a variety of comorbidities consistent with patients that receive both echocardiography and abdominal ultrasound studies with prevalent hypertension (28.8%), hyperlipidemia (20.1%), diabetes (19.3%), hepatitis B (1.2%) and hepatitis C (3.7%) (**Table 1**). The average BMI was 26.5± 6.2, and 5.3% of the patients regularly consumed alcohol. The median duration between the echocardiography and abdominal ultrasound examinations was 0 days (interquartile range, -4 to +3 days). In this cohort, there were 371 (8.8%) cirrhosis and 645 (14.0%) SLD cases based on abdominal ultrasound reports.

**Table 1:** Patient characteristics of the study cohort.

| Characteristic | CSMC cohort,<br>N = 24,276 | CSMC Ultrasound<br>Test cohort<br>N = 1,486 | CSMC MRI Test<br>cohort,<br>N = 2,335 | SHC External<br>Validation cohort,<br>N = 106 |
| --- | --- | --- | --- | --- |
| Number of studies | 66,922 | 6,996 | 6,751 | 106 |
| Number of video files | 1,596,640 | 163,736 | 164,579 | 5,388 |
| Age, mean (SD) | 65.1 (17.1) | 63.0 (17.3) | 62.7 (14.6) | 59.3 (16.8) |
| BMI, mean (SD) | 27.1 (6.6) | 26.9 (6.6) | 26.9 (6.2) | 27.0 (7.9) |
| Gender, female | 11,892 (49.1%) | 666 (44.9%) | 1,200 (51.4%) | 48 (45.3%) |
| Race/Ethnicity |  |  |  |  |
| White | 16,347 (67.3%) | 972 (65.4%) | 1,572 (67.3%) | 49 (46.2%) |
| Black | 3,962 (16.3%) | 279 (18.8%) | 335 (14.3%) | 2 (1.9%) |
| Asian | 1,699 (7.0%) | 95 (6.4%) | 184 (7.9%) | 21 (19.8%) |
| Other | 2,268 (9.3%) | 140 (9.4%) | 244 (10.5%) | 34 (32.0%) |
| Hypertension | 6,989 (28.8%) | 454 (30.6%) | 549 (23.5%) | 72 (67.9%) |
| Dyslipidemia | 4,879 (20.1%) | 328 (22.1%) | 367 (15.7%) | 45 (42.5%) |
| Diabetes | 4,675 (19.3%) | 310 (20.9%) | 449 (19.2%) | 43 (40.6%) |
| Stroke | 1,626 (6.7%) | 113 (7.6%) | 107 (4.6%) | 7 (6.6%) |
| Atrial Fibrillation | 802 (3.3%) | 71 (4.8%) | 42 (1.8%) | 36 (34.0%) |
| Heart Failure | 2,964 (12.2%) | 276 (18.6%) | 105 (4.5%) | 47 (44.3%) |
| Coronary Artery<br>Disease | 4,517 (18.6%) | 342 (23.0%) | 259 (11.1%) | 29 (27.4%) |
| LVEF, mean (SD) | 58.1 (14.9) | 56.5 (16.5) | 63.0 (10.1) | 55.4 (13.8) |
| Active Smoking | 1,208 (5.0%) | 100 (6.7%) | 101 (4.3%) | 4 (3.8%) |
| HBV | 284 (1.2%) | 15 (1.0%) | 72 (3.1%) | 9 (8.5%) |
| HCV | 908 (3.7%) | 53 (3.6%) | 201 (8.6%) | 9 (8.5%) |
| Alcohol | 1,243 (5.1%) | 92 (6.2%) | 211 (9.0%) | 26 (24.5%) |

LVEF: Left Ventricular Ejection Fraction; BMI: Body Mass Index; SD: Standard Deviation; SLD: Steatotic liver disease, HBV: hepatitis B, HCV: hepatitis C

We identified an additional test population of 6,959 echocardiogram videos from 4,145 studies across 2,335 CSMC patients who underwent abdominal MRI for cirrhosis and MRI with PDFF/MRS for SLD, and echocardiography within 365 days. The patients in this additional test cohort were not included in training and validation cohort of disease detection models. In the SHC cohort for EchoNet-Liver evaluation, we identified 66 studies of individual patients who have a total of 130 high-quality subcostal videos. Of all patients, 10 and 11 patients were diagnosed with cirrhosis and SLD respectively based on abdominal ultrasound. Patient characteristics are presented in **Table 1**. Patient characteristics in each model development (i.e., view-classifier model, quality-control model, cirrhosis detection model and SLD detection model) were demonstrated in **Supplemental Tables 1-4**.

### View Classifier and Quality Control Performance Across Two Institutions

A total 11,419 subcostal view videos were used for training a subcostal view classifier model, with all other videos labeled as non-subcostal controls. On an independent test set of 100 CSMC studies (n = 2,315 videos), the view classifier model identified 186 out of 196 subcostal view videos with an AUC of 0.991 (0.984 – 0.997). The sensitivity was 0.949 (0.910-0.974) and specificity was 0.999 (0.998-1.00). In the SHC population, the subcostal view classifier identified 149 out of 228 subcostal videos from a total of 5,280 total videos with an AUC of 0.965 (0.956 - 0.974). The sensitivity was 0.654 (0.591 – 0.716) and specificity was 0.993 (0.991 – 0.995). In comparison of image quality by two cardiologists, the quality-control model demonstrated an AUC of 0.855 (0.800-0.905) in CSMC and an AUC of 0.785 (0.722-0.843) in SHC.

### Disease Detection Performance Across Two Institutions

In the held-out CSMC test ultrasound dataset, the EchoNet-Liver detected chronic liver disease with an AUC of 0.837 (95% CI 0.789 - 0.880) for cirrhosis and 0.799 (0.758 - 0.837) for SLD (**Figure 2**). For cirrhosis, the AUPRC was 0.309 (0.206 – 0.417), PPV was 0.238 (0.180 - 0.299), and NPV was 0.976 (0.965 - 0.986). For SLD, the AUPRC was 0.408 (0.325 – 0.491), the PPV was 0.275 (0.232 - 0.319) and the NPV was 0.951 (0.935 - 0.966). In the CSMC test cohort, the algorithm showed a sensitivity of 0.696 (0.582 - 0.803) and a specificity of 0.847 (0.824 - 0.868) for detecting of cirrhosis and sensitivity of 0.741 (0.669 - 0.812) and specificity of 0.720 (0.692 - 0.747) for detecting SLD. In the SHC cohort, EchoNet-Liver detected cirrhosis and SLD with an AUC of 0.830 (0.738 - 0.909) and 0.768 (0.652 – 0.875) respectively. For cirrhosis, the AUPRC was 0.500 (0.278 – 0.708), PPV was 0.334 (0.203 - 0.471), and NPV was 0.952 (0.901 - 0.989). For SLD, the AUPRC was 0.518 (0.299 – 0.705), the PPV was 0.370 (0.219 - 0.528) and the NPV was 0.924 (0.864 - 0.976). In the SHC test cohort, EchoNet-Liver demonstrated a sensitivity of 0.802 (0.611 - 0.957) and a specificity of 0.709 (0.623 - 0.789) for detecting of cirrhosis and sensitivity of 0.667 (0.450 - 0.867) and specificity of 0.781 (0.701 - 0.855) for detecting SLD. Evaluation metrics are summarized in **Table 2**.

**Figure 2.**
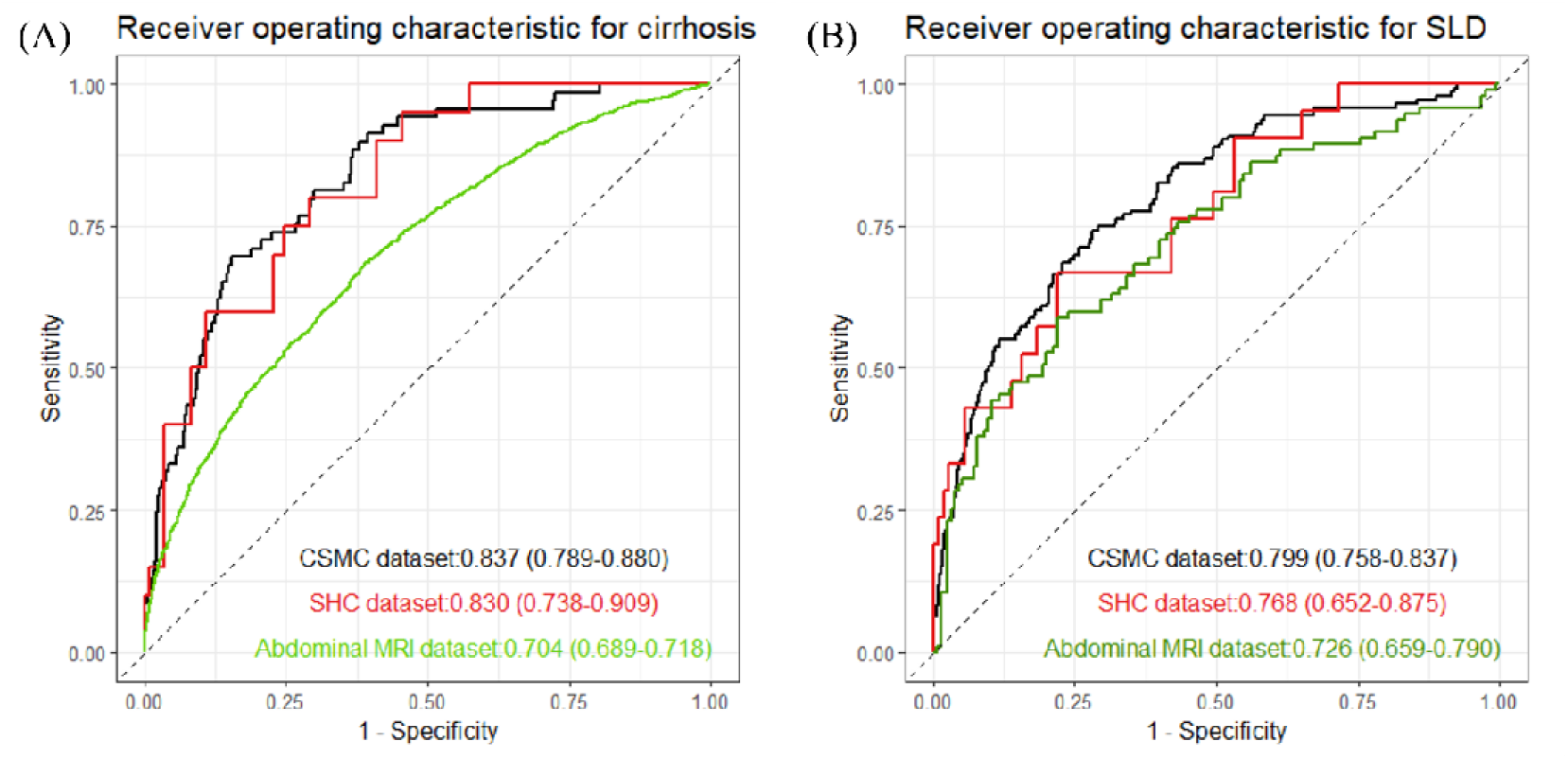
Model Performance of Echo-Net-Liver. Performance of a deep learning model using high-quality subcostal echocardiography videos for cirrhosis (A) and SLD (B). Model was evaluated in an internal CSMC held-out test dataset (black), external SHC abdominal ultrasound dataset (red) and abdominal MRI test dataset (green). US: ultrasound, MRI: Magnetic resonance imaging, SLD: Steatotic liver disease

**Table 2:** Prediction of liver disease by deep learning analysis of echocardiography using labels from abdominal ultrasound held-out test population.

|  | AUROC | AUPRC | Sensitivity | Specificity | PPV | NPV |
| --- | --- | --- | --- | --- | --- | --- |
| <u>(a) CSMC held-out test cohort</u> |  |  |  |  |  |  |
| Cirrhosis | 0.837<br>(0.789 - 0.880) | 0.309<br>(0.206 – 0.417) | 0.696<br>(0.582 - 0.803) | 0.847<br>(0.824 - 0.868) | 0.238<br>(0.180 - 0.299) | 0.976<br>(0.965 - 0.986) |
| SLD | 0.799<br>(0.758 - 0.837) | 0.408<br>(0.325 – 0.491) | 0.741<br>(0.669 - 0.812) | 0.720<br>(0.692 - 0.747) | 0.275<br>(0.232 - 0.319) | 0.951<br>(0.935 - 0.966) |
| <u>(b) SHC external validation test cohort</u> |  |  |  |  |  |  |
| Cirrhosis | 0.830<br>(0.738 - 0.909) | 0.500<br>(0.278 – 0.708) | 0.802<br>(0.611 - 0.957) | 0.709<br>(0.623 - 0.789) | 0.334<br>(0.203 - 0.471) | 0.952<br>(0.901 - 0.989) |
| SLD | 0.768<br>(0.652 - 0.875) | 0.518<br>(0.299 – 0.705) | 0.667<br>(0.450 - 0.867) | 0.781<br>(0.701 - 0.855) | 0.370<br>(0.219 - 0.528) | 0.924<br>(0.864 - 0.976) |

PPV: positive predictive value. NPV: positive predictive value. AUROC: Area under receiver operating characteristic. SLD: Steatotic liver disease.

### Comparison with Diagnosis by Magnetic Resonance Imaging

In order to evaluate the performance of EchoNet-Liver across multiple diagnostic pathways, the algorithm was evaluated in a cohort of patients that both received echocardiography and abdominal MRI at CSMC. In the MRI paired cohort, EchoNet-Liver detected the presence of cirrhosis with an AUC of 0.704 (95% CI 0.689–0.718) (**Figure 2-A**). and a AUPRC of 0.493 (95% CI 0.468–0.518) (**Supplementary Figure 2-A)**. When evaluated in the MRS/PDFF steatosis cohort, the model detected the presence of SLD with an AUC of 0.726 (95% CI 0.659–0.790) (**Figure 2-B**) and an AUPRC of 0.626 (95%CI 0.519–0.731) (**Supplementary Figure 2-B)**

### Model Explainability

We generated saliency maps for representative echocardiography images from the two test datasets (CSMC held-out dataset and SHC external dataset shown in **Figure 3**). For both cirrhosis and SLD, EchoNet-Liver highlighted the liver of the subcostal echocardiographic images as regions of interest, with a diffuse activation throughout the hepatic parenchyma.

**Figure 3:**
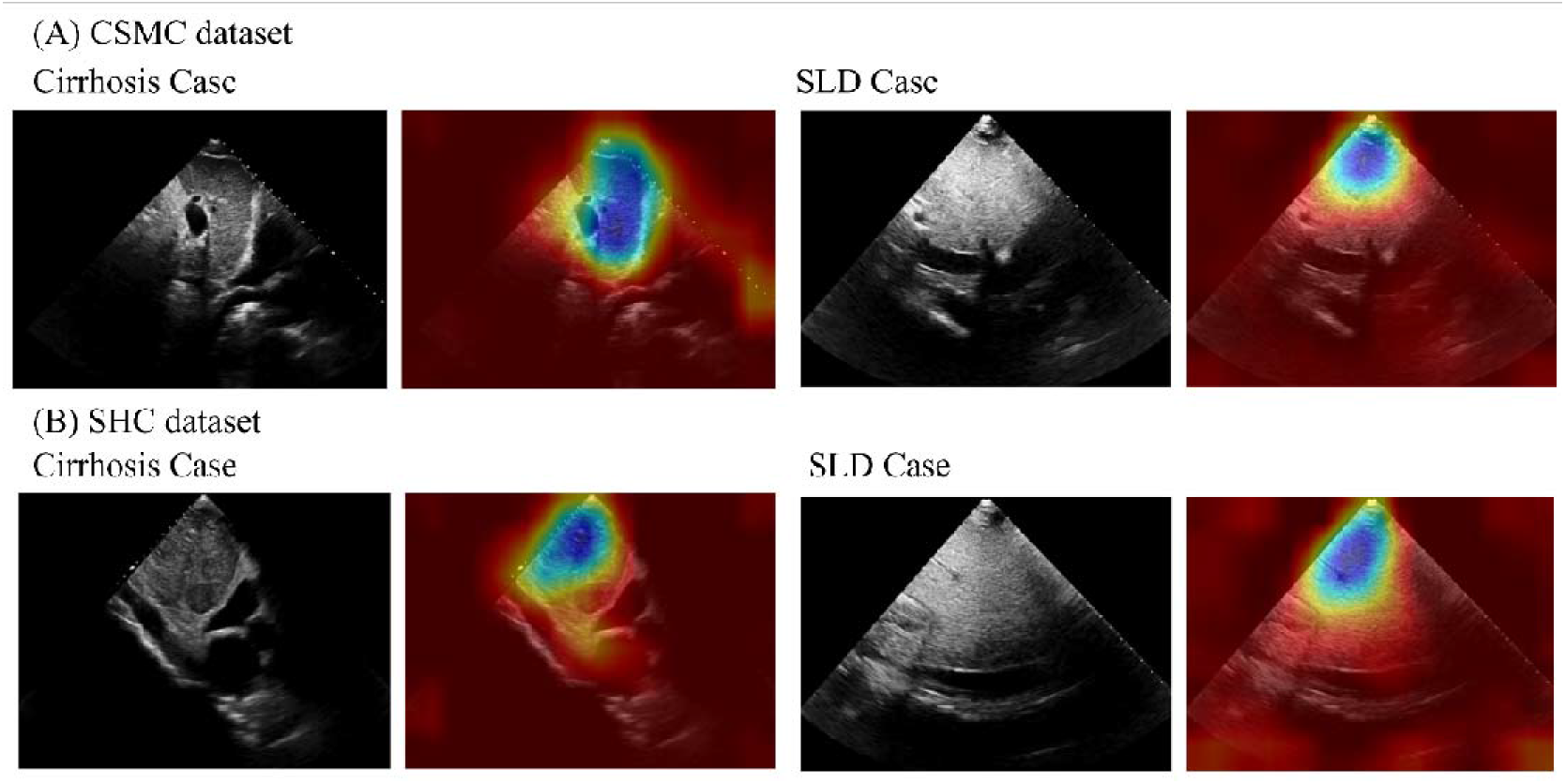
Representative saliency maps for the two test datasets. Corresponding pairs of input echocardiogram frames and Grad-CAM visualization of both cirrhosis and steatotic liver disease (SLD) (CSMC dataset (A) and Stanford Healthcare dataset (B)) Pixels brighter in color and closer to blue were more salient to model predictions, while those darker in color were less important to the model’s final prediction.

## Discussion

In this study, we demonstrated strong performance of a deep learning pipeline (EchoNet-Liver) for detecting cirrhosis and SLD from clinical echocardiography images. The discriminative ability of the model was confirmed in a geographically distinct external health-care system cohort as well as in a cohort of patients with paired abdominal MRI imaging. Across diverse populations and disease definitions, deep learning-enhanced echocardiography enabled high-throughput automated detection of chronic liver disease, which could enable opportunistic screening for asymptomatic liver disease.

Chronic liver disease often remains undiagnosed due to the asymptomatic nature of early disease. Despite the significant prevalence and morbidity, routine screening is not recommended given the high cost of imaging and lack of evidence of its cost-effectiveness^23^. Opportunistic screening from echocardiogram images can identify a high-risk population of patients with concurrent cardiovascular risk and liver disease in a cost-effective manner. By harnessing pre-existing imaging indicated for other diagnostic reason, our AI-enhanced workflow can increase the potential utility of imaging examinations^29^. By incorporating view classification, quality control, and disease detection in one pipeline, automation with AI can enable high-throughput evaluation of this non-invasive and common cardiovascular diagnostic test.

There are several limitations in the present study to consider. This study is a retrospective study conducted at two tertiary care centers for patients who have undergone both abdominal ultrasound and echocardiography, which may result in selection bias. The spectrum of age, gender, race, and comorbidities in the study dataset may not represent the general population and may bias towards patients with more comorbidity, necessitating further external validation. Comparison between disparate imaging modalities such as ultrasound and MRI will have inherent limitations due to different modality-specific accuracy. However, in totality this study suggests that the clinical utility of high throughput disease screening using AI is promising, particularly for early disease, and enhances the utility of pre-existing imaging data^11,12,30^. Further studies are warranted to establish the optimal clinical workflow for opportunistic liver disease screening among CVD patients and downstream treatment. By improving diagnosis of subclinical CLD, we may be able to limit or reverse disease progression ^20–22^ and improve patient care by triaging patients toward appropriate clinical and diagnostic management^23,24^.

In conclusion, we found that EchoNet-Liver, a deep learning pipeline using echocardiography to detect the presence of SLD and cirrhosis, had strong performance in multiple populations and disease definitions. The findings were consistent across two institutions and with comparison to abdominal magnetic resonance imaging. Deep learning applied to echocardiography may offer an opportunity for opportunistic and cost-effective screening for chronic liver disease.

## Author contributions

Concept and design: YS, DO, AK

Code and programming: YS, MV, JR

Acquisition, analysis, or interpretation of data: YS, DO, AK, FA

Drafting and Critical revision of the manuscript: YS, DO, AK, HT, SC JC

Statistical analysis: YS, DO, AK

Obtained funding: AK, DO, SC

Supervision: AK, DO, SC

## Abbreviations

AI: Artificial intelligence
AUROC: Area under receiver operating characteristic
AUPRC: Area Under precision recall curve
CLD: chronic liver disease
CVD: cardiovascular disease
LVEF: Left ventricular ejection fraction
PR: Precision-recall
SLD: steatotic liver disease
PPV: positive predictive value
NPV: negative predictive value
MRI: magnetic resonance imaging
MRS: magnetic resonance spectroscopy
PDFF: proton density fat fraction

